# Toward a unified classification of acute myeloid leukemia, myelodysplasia-related: a multicenter retrospective study

**DOI:** 10.64898/2026.06.15.26355739

**Authors:** Ying Liu, Derek M. Loneman, Brenden Bready, David Nemirovsky, Alexa Cohen, Xin Wang, Eytan Stein, Yanming Zhang, Andriy Derkach, Robert P. Hasserjian, Wenbin Xiao

## Abstract

**Key points:**

- AML-MR is prognostically distinct from AML with *TP53* mutations.
- AML-MR is unified by MR gene mutations and MR cytogenetic abnormalities; isolated *RUNX1* mutations, trisomy 8, and del(20q) may warrant exclusion.

The 5th Edition of the World Health Organization Classification of Haematolymphoid Neoplasms (WHO5th) and the 2022 International Consensus Classification (ICC) both recognize myelodysplasia-related acute myeloid leukemia (AML-MR) as a diagnostic entity increasingly defined by integrated genomic data. Although largely concordant, the two classifications differ in various ways that should be resolved to achieve future harmonization. To address the areas of uncertainty, we retrospectively analyzed 615 newly diagnosed AML cases from adult patients treated at two large cancer centers. We demonstrate that AML-MR, whether defined by gene mutations (MR-GM) or cytogenetic abnormalities (MR-CGA), constitutes a prognostically distinct group with inferior outcome compared to most AML subtypes, second only to *TP53*-mutated or *EVI1*-rearranged AML. Isolated *RUNX1* mutations were not associated with antecedent myeloid neoplasia. Neither the number of mutated MR genes nor their variant allele frequency independently impacted outcomes. Trisomy 8 and del(20q) did not confer inferior outcomes and may warrant exclusion from MR-CGA. Complex karyotype without *TP53* mutations did not worsen outcomes within AML-MR and may be considered equivalent to other MR-CGA. The adverse prognosis of AML-MR appeared to be at least partly driven by *ASXL1* and/or *EZH2* mutations. These findings provide evidence toward a unified schema across the WHO5th and ICC.

## Introduction

Accurate classification of acute myeloid leukemia (AML) is essential for diagnosis, prognostication, and treatment selection, and has evolved substantially since the initial codification of AML subtypes in the 3rd edition of WHO Classification in 2003. Current classification schemas rely increasingly on cytogenetic and molecular findings as the best available approximation of the underlying leukemic biology. This paradigm shift from morphologic to genetic classification is reflected in both the WHO5th and ICC classification systems,^1,2^ which exhibit significant overlap but also important differences in several areas. In parallel, risk-stratification frameworks, most prominently those developed by the European Leukemia Network (ELN),^3,4^ have adopted an ever-greater reliance on genetic data to guide clinical management. Despite this progress, disagreement persists regarding the prognostic significance of certain genetic findings, alone or in combination, particularly in the context of AML-MR.

Both WHO5th and ICC recognize AML-MR as a distinct diagnostic category, defined predominantly by integrated molecular genetic changes.^1,2^ The diagnostic criteria substantially overlap between the two systems but diverge in several clinically meaningful ways that have created confusion for pathologists and clinicians.^5^ Both systems recognize 8 canonical mutations originally delineated as myelodysplastic syndrome/neoplasm (MDS)-associated by Lindsley and colleagues based on their association with secondary AML progressed from known MDS.^6^ There are 8 mutations - *ASXL1*, *BCOR*, *EZH2*, *SF3B1*, *SRSF2*, *STAG2*, *U2AF1*, and *ZRSR2 –* are considered AML-MR defining. Notably, however, the ICC additionally includes *RUNX1* mutations as AML-MR-defining based on their reported association with AML arising from antecedent myeloid neoplasia (AMN) and their potential to confer adverse outcomes.^7^ Because *RUNX1* mutations frequently co-occur with other MR gene mutations, it remains unclear whether these associations represent “guilt by association” or whether isolated *RUNX1* mutations genuinely confer a biology similar to other canonical MR mutations.^6,8–13^ Although not incorporated into either classification scheme, some studies have suggested that the number or variant allele frequency (VAF) of MR-associated mutation may impact their prognostic relevance.^11,14^

A second area of divergence concerns the role of clinical ontogeny. The WHO5th retains prior history of MDS or MDS/myeloproliferative neoplasm (MDS/MPN) as a diagnostic criterion for AML-MR, regardless of the underlying genetic profile. In contrast, the ICC treats ontogeny as a diagnostic qualifier rather than a primary defining criterion. Although a recent study by McCarter, et al demonstrated that AML ontogeny has independent prognostic value across all AML subtypes,^15^ the diagnostic relevance of clinical ontogeny in the molecular era requires further evaluation.^16^

Third, whereas the WHO5th considers MR gene mutations (GM) and cytogenetic aberrations (CGA) as equivalent for classification purposes, the ICC subdivides AML-MR into two groups defined by GM or CGA, with MR-GM superseding MR-CGA in a hierarchical classification scheme. Whether this prioritization carries prognostic or biological significance has not been rigorously validated in large cohorts.

Finally, given increasing evidence for unique biology characterized by frequent bi-allelic inactivation, complex karyotype (CK), and dismal outcomes,^17^ AML with mutated *TP53* is now recognized as a distinct entity within the ICC, superseding other categories, including AML-MR, in which *TP53* mutations are frequently encountered. Whether *TP53*-mutated AML is truly prognostically distinct from other AML-MR subgroups, and whether complex karyotype in the absence of *TP53* mutations worsens prognosis within AML-MR, remain open to questions.

In this retrospective, multi-institutional cohort study, we sought to address several unresolved questions in AML-MR classification: (1). Do isolated *RUNX1* mutations, in the absence of the 8 canonical MR mutations shared by ICC and WHO, associate with AMN and/or adverse prognosis? (2). Does AML-MR defined by MR-GM versus MR-CGA exhibit clinically meaningful differences in outcome? (3). Does the number or VAF of MR-GMs impact prognosis? (4). Does CK, independent of *TP53* mutation, confer additional adverse risk within AML-MR? (5). Is the prognosis of *TP53-*mutated AML, as defined by ICC, truly distinct from AML-MR defined by GM and CGA? To investigate these questions, we analyzed 615 newly diagnosed AML cases with comprehensive molecular, cytogenetic, and clinical annotation from two major cancer centers. Our findings offer additional clarity to support a refined, unified definition of AML-MR.

## Method and Material

### Study cohort

A total of 615 adult patients with newly diagnosed AML (excluding acute promyelocytic leukemia) were retrospectively identified from two academic medical centers: Memorial Sloan Kettering Cancer Center (MSKCC; n=404) and Massachusetts General Hospital (MGH; n=211), spanning the period from 2015 to 2025 (**Fig. 1A**). All patients underwent comprehensive cytogenetic evaluation, including conventional karyotyping and fluorescence in situ hybridization (FISH), as well as mutational profiling by targeted next-generation sequencing (NGS). Clinical, histopathological, and molecular characteristics were manually curated, and all cases were classified according to both WHO5th and ICC.

**Figure 1:**
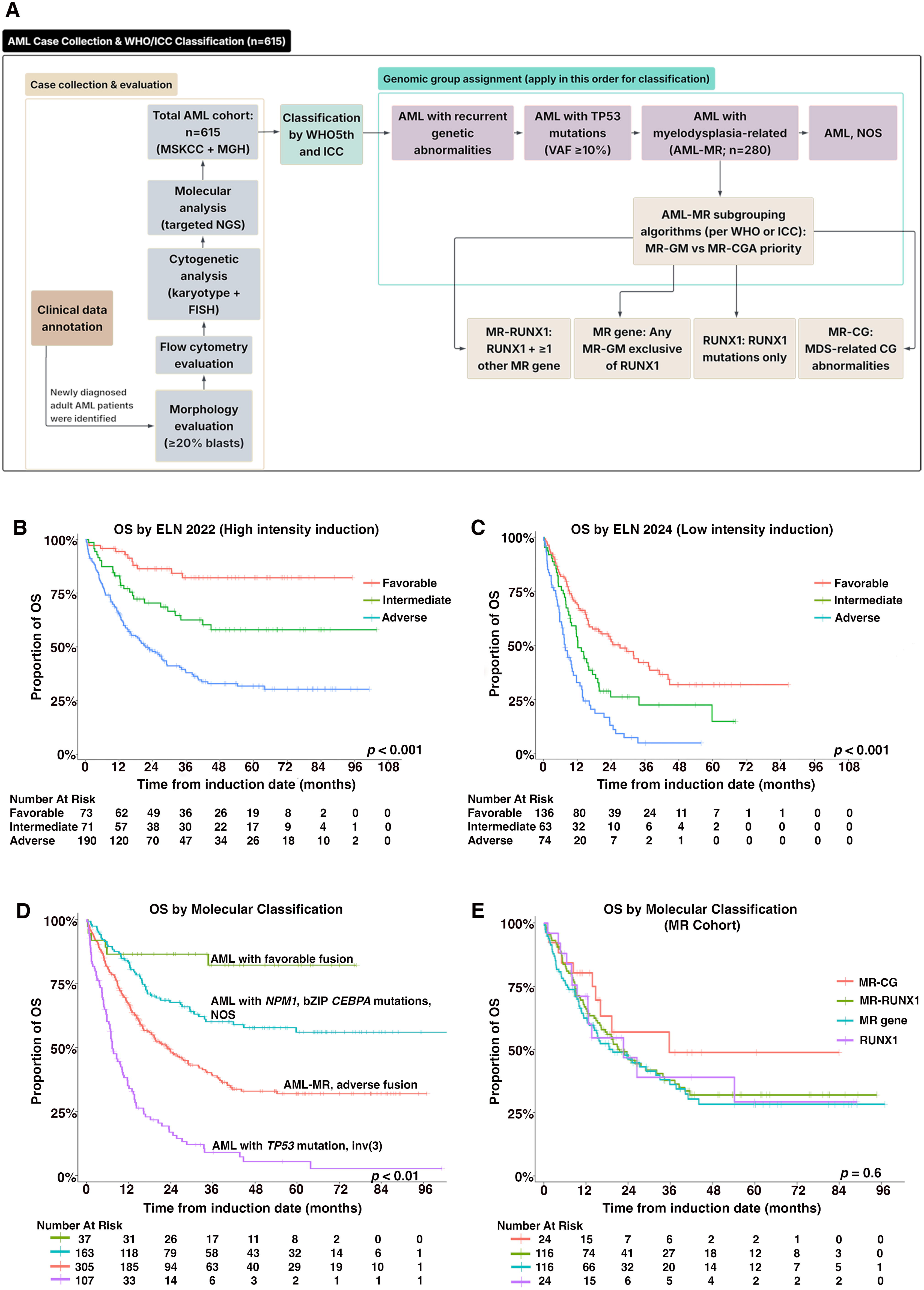
Study overview, cohort validation, and molecular classification. **(A)** Schematic of case collection, genomic annotation, and diagnostic classification across two institutions (MSK and MGH). **(B)** Overall survival stratified by ELN 2022 risk groups (favorable, intermediate, and adverse) in patients treated with high-intensity induction. **(C)** Overall survival stratified by ELN 2024 risk groups in patients treated with low-intensity induction. **(D)** Overall survival of all patients classified into four major molecular subgroups according to WHO5th and ICC. **(E)** Overall survival of four AML-MR subgroups defined by ICC, with mutation-based criteria (MR-GM) prioritized over cytogenetic criteria (MR-CGA).

AML-MR was defined by the presence of MR-GM and/or MR-CGA as specified by either classification system. Notably, AML cases harboring *TP53* mutations were excluded from the AML-MR category in this analysis. Additionally, cases with antecedent MDS or MDS/MPN in the absence of concurrent MR-GM and/or MR-CGA were excluded from the AML-MR cohort and analyzed as a separate group. AML-MR cases were subsequently subdivided into four groups based on the presence or absence of MR-GM and MR-CGA:

(1). RUNX1*: RUNX1* mutation in the absence of any of the 8 canonical MR mutations;
(2). MR gene: mutation in at least 1 of the 8 canonical MR genes (excluding *RUNX*1);
(3). RUNX1-MR: mutations in *RUNX1* and at least 1 of the 8 canonical MR genes;
(4). MR-CG: MR defining cytogenetic abnormality.

To account for the frequent co-occurrence of MR-GM and MR-CGA, and to address whether prioritization of MR-GM versus MR-CGA is relevant to classification or prognostication, four classification algorithms were employed:

(1). Prioritizing MR-GM over CGA as defined by ICC;
(2). Prioritizing MR-CGA over MR-GM as defined by ICC;
(3). Prioritizing MR-GM over CGA as defined by WHO5th;
(4). Prioritizing MR-CGA over MR-GM as defined by WHO5th.

For other AML diagnostic groups (Table 1), the following definitions were applied: Fusion-favorable includes AML with t(8;21), or inv(16). Fusion-adverse includes AML with t(6;9) and *KMT2A* rearrangements but excluding *EVI1* rearrangements; all cases with *KMT2A* rearrangement were included in the fusion-adverse group to facilitate statistical analysis given limited case numbers of AML with t(9;11). *NPM1* includes AML with mutated *NPM1*. *CEBPA* includes AML with in-frame bZIP *CEBPA* mutations. *TP53* includes AML with at least 1 *TP53* mutation with a VAF ≥10%, per ICC criteria; multi-hit versus single-hit status was determined for all *TP53*-mutated cases. All cases were stratified by ELN 2022 risk criteria if treated with intensive chemotherapy (7+3 or equivalent regimens or Vyxeos) and by ELN 2024 risk criteria if treated with non-intensive regimens (azacitidine+venetoclax). ECOG performance status (PS) was determined at the time of AML diagnosis.

**Table 1:**
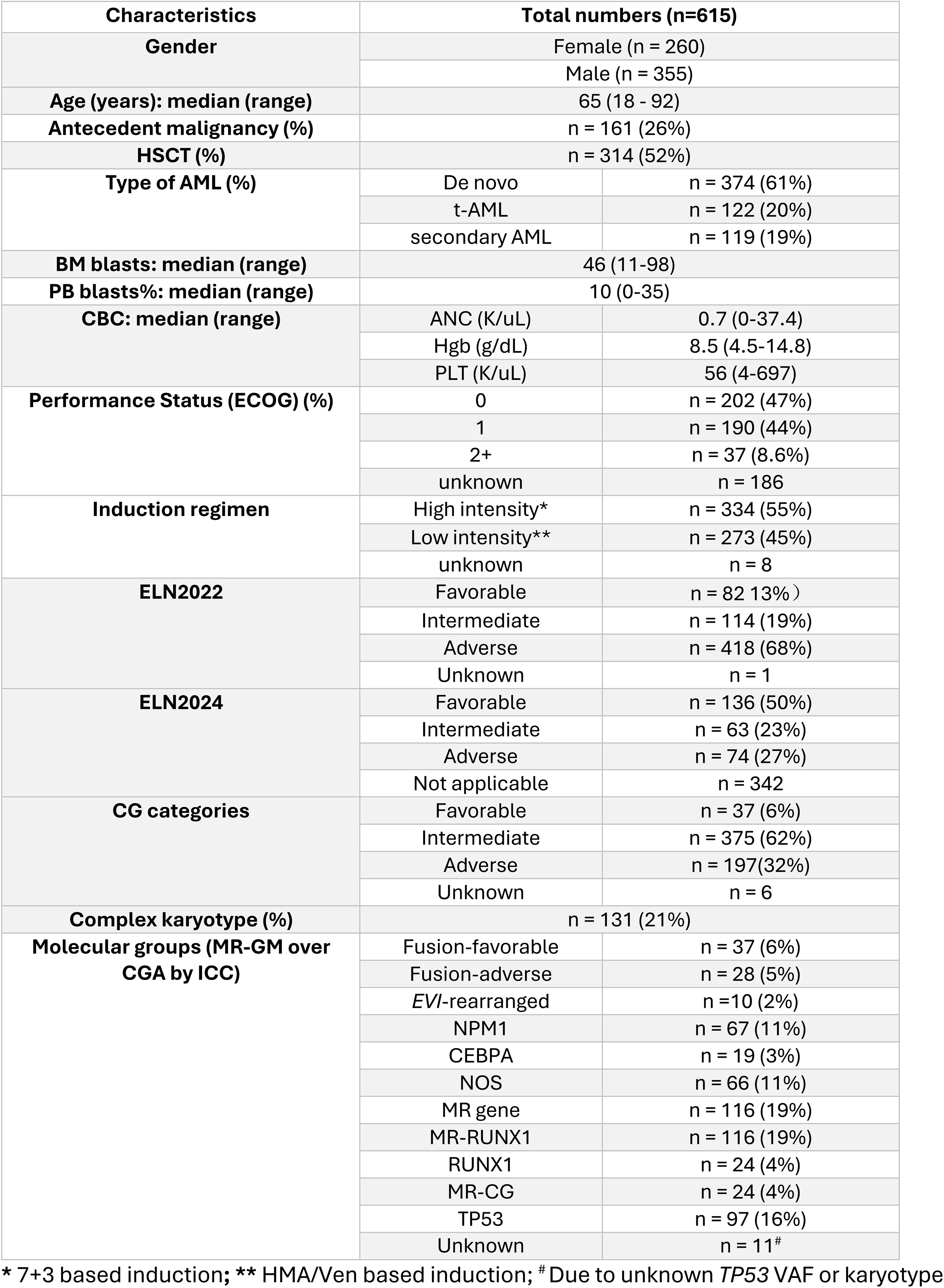
Clinicopathologic characteristics of all patients.

### Targeted next-generation sequencing

Mutational profiling was performed on bone marrow aspirate or peripheral blood obtained at diagnosis using institution-specific NGS platforms. At MSKCC, profiling was performed using MSK-IMPACT-HEME (MSK Integrated Mutational Profiling of Actionable Cancer Targets), a custom hybridization capture-based NGS assay detecting somatic mutations and copy number alterations in coding regions of at least 400 genes.^18^ Library preparation, sequencing, variant calling, and annotation were performed using validated pipelines as previously described.^19^ Variant calling was performed in paired-sample mode with manual curation alongside corresponding matched normal (nail and donor) samples.

At MGH, samples were profiled by either the Heme Snapshot or Rapid Heme Panel assays. Heme Snapshot is a DNA-based, anchored multiplex polymerase chain reaction assay designed to identify single nucleotide variants and insertions/deletions across numerous genes associated with hematologic malignancy. ^20^ The Rapid Heme Panel is a custom amplicon-based NGS panel covering mutational hotspots in oncogenic driver genes and whole coding regions of tumor suppressor genes associated with hematologic neoplasia.^21^

Multi-hit status for *TP53* mutation was defined as the presence of (1) ≥2 distinct *TP53* mutations with a VAF of at least one mutation ≥10%; or (2) a single *TP53* mutation associated with at least one of the following: deletion involving the *TP53* locus at 17p13.1 evident on cytogenetic analysis, VAF ≥ 50%, or copy neutral loss of heterozygosity (CN-LOH) at the *TP53* locus on 17p.

### Cytogenetic analysis

Conventional chromosome analysis of bone marrow aspirate and peripheral blood was performed following standard procedures with analysis of at least 20 metaphase cells per case. Chromosome abnormalities were recorded per the International System for Human Cytogenetic Nomenclature (2016). At MSKCC, FISH analysis was routinely performed for recurring chromosomal abnormalities in AML, including *KMT2A* (11q23), *MECOM* (3q26.2), deletion 5q31, or loss of chromosome 5, trisomy 8, deletion 7q31, or loss of chromosome 7, deletion 17p13.1 (TP53), or loss of chromosome 17. All FISH probes were from Abbott Molecular (Des Plaines, IL) and validated in the laboratory. At MGH, FISH analysis was performed on selected cases as deemed necessary at the time of diagnosis (e.g., to assess deletion of the *TP53* locus on 17p13.1). All cytogenetic results were re-annotated by a board-certified cytogeneticist. Cytogenetic risk groups (favorable, intermediate, adverse) were assigned based on 2022 ELN criteria.

### Statistical analysis

Descriptive statistics are reported as median and interquartile range (IQR) for continuous variables and as counts and percentages for categorical variables. Association between categorical variables was assessed using the Fisher exact test or χ2 test, as appropriate. Differences in a continuous variable between and among patient groups were evaluated using the Wilcoxon rank-sum test. Overall survival (OS) was calculated from the date of induction therapy to the date of death or last follow up. The Kaplan-Meier method was used to estimate OS distributions, and differences between groups were assessed using the log-rank test. Univariable Cox proportional hazard regression was performed to evaluate OS association with genetic and clinical characteristics. To adjust these for demographic and clinical characteristics, age, gender, CK, type of AML, transplant (modeled as a time-dependent variable), and ECOG PS were assessed in multivariable Cox-regression models which were stratified by treatment and center. All analyses and graphics were produced using R version 4.5.1.

## Results

### Cohort characteristics and validation of ELN risk stratification

Clinicopathologic characteristics of the cohort are summarized in **Table 1**. A total of 615 adult patients were included, with a median age of 65 years (range: 18-92 years) and a male-to-female ratio of 3:2. De novo, therapy-related, and secondary AML (arising from antecedent MDS and/or MDS/MPN but excluding therapy-related) accounted for 61%, 20%, and 19% of all patients, respectively; 30% of therapy-related AML patients were progressed from therapy-related MDS or MDS/MPN. Allogeneic stem cell transplant (ASCT) was performed in 314 (52%) patients.

To validate the quality of cohort annotation and clinical data, ELN risk stratification was applied separately by treatment intensity. Among the 334 (55%) patients receiving high-intensity induction chemotherapy, ELN 2022 risk stratification significantly stratified favorable, intermediate and adverse risk groups (median OS: unreached, unreached, and 22 months, respectively; *p*<0.001). Among the 273 (45%) patients receiving low-intensity induction chemotherapies, ELN 2024 significantly stratified favorable, intermediate and adverse risk groups (median OS: 27, 12, and 7.6 months, respectively; *p*<0.001) (**Fig. 1B-C**), whereas ELN 2022 demonstrated less optimal separation this subset (data not shown). These data confirm the validity of cohort annotation and support the use of ELN 2024 for patients receiving non-intensive chemotherapy.

The entire cohort was then classified by molecular subtypes, and Kaplan-Meier analysis for OS was performed. AML with favorable fusions had the most favorable outcome (median OS: unreached), followed by *CEBPA* (median OS: unreached) and *NPM1* categories (median OS: unreached). AML with *TP53* mutations (median OS: 6.9 months) or *EVI*1-rearrangement (median OS: 14 months) showed the shortest survival. AML-MR cases (median OS: 23, 20, 22 and 36 months for RUNX1, MR gene, MR-RUNX1, and MR-CG, respectively), in together with AML with adverse fusion (including t(6;9) and *KMT2A* rearrangements), showed an outcome (median OS: 23 months) that was inferior to most AML groups, only second to *TP53* mutations (≥1 *TP53* mutation with ≥10% VAF) and *EVI*1-rearrangement (median OS: 7.3 months) (*p*<0.001) (**Fig. S1**). To simplify data visualization, individual molecular classes collapsed into a total of 4 larger groups based on similar OS (**Fig. 1D**).

Univariable analysis identified age, AML ontogeny (de novo, secondary, or therapy-related), CG risk groups by ELN 2022, CK, ECOG PS, treatment (high-intensity versus low-intensity induction) and ASCT as significant prognostic factors for the entire cohort (**Table S1**). In multivariable analysis, after stratifying treatment intensity and institution (MSKCC versus MGH) the following factors retained independent prognosis: sex, AML ontogeny, CK, ECOG PS, and ASCT (**Table S2**).

### AML-MR defined by GM and/or CGA form a prognostically distinct group

Of the 615 patients, 280 (45.5%) met diagnostic criteria by WHO5th and/or ICC for AML-MR (**Table 1, Table S3**). When MR-GM was prioritized over MR-CGA by ICC criteria, used here as the representative classification algorithm, the majority of AML-MR patients were classified on the basis of MR-GM: 19% of the total cohort had mutations in at least one of the 8 canonical MR genes with concurrent *RUNX1* mutation (RUNX1-MR), and 19% had mutations in at least one MR gene without *RUNX1* mutation (MR gene). Patients meeting ICC criteria based on isolated *RUNX1* mutation alone (RUNX1) or MR-CGA in the absence of MR-GM (MR-CG) each comprised 4% of the total cohort.

Regardless of whether WHO5th or ICC criteria were applied, or whether MR-GM or MR-CGA was prioritized in classification, the four AML-MR subgroups (RUNX1, RUNX1-MR, MR gene, and MR-CG) consistently formed a discrete prognostic group among all AML cases (**Fig. S1A-D**). No statistically significant difference in OS was observed among the four AML-MR subgroups (median OS: 23, 22, 20, and 36 months for RUNX1, RUNX1-MR, MR gene, and MR-CG, respectively; *p*=0.6), regardless of classification schema or GM/CGA prioritization (**Fig. 1E, Fig. S2**).

### Isolated *RUNX1* mutations are not associated with antecedent myeloid neoplasia

To further characterize the clinical and molecular features of isolated *RUNX1*-mutated AML, we analyzed ontogeny, prior therapy exposure, and clonal architecture across AML-MR subgroups. Only 1/24 (4%) of patients in the RUNX1 subgroup had a history of AMN, whereas 43/116 (37%, *p*=0.003) of MR-RUNX1 patients and 32/116 (28%, *p*=0.03) of MR patients had antecedent disease history (**Table S4**). Conversely, RUNX1 patients more frequently had a prior history of cytotoxic therapy exposure (8/24, 33%) compared to RUNX1-MR (13/116, 11%, *p*=0.1) or MR gene (12/116, 10%, *p*=0.04) patients (**Table S4**).

When MR-CGA was prioritized over MR-GM to identify truly isolated *RUNX1* mutations (i.e. without concurrent MR-CGA or MR-GM), patients in *RUNX1* group demonstrated numerically superior OS compared to all other AML-MR groups (median OS: 54 months vs 20 months; *p*=0.2), although this difference did not reach statistical significance, likely due to the small number of cases (n=12) (**Fig. S3**).

To further investigate the clonal role of *RUNX1* mutations, hierarchical VAF analysis was performed in the RUNX1-MR subgroup by comparing the VAF of *RUNX1* to co-occurring MR-GMs. After exclusion of X-linked genes (*STAG2*, *BCOR*, and *ZRSR2*), the median VAF of dominant MR-GMs was 41.4% (IQR 35-47%), significantly higher than that of *RUNX1* mutations (39.5%; IQR 26-47%, *p*=0.03, **Fig. S4**).

Correlation analysis across the entire cohort confirmed significant positive correlation between *RUNX1* mutations and all other MR-GMs except *ZRSR2* (likely due to limited numbers), as well as its significant negative correlation between *RUNX1* and *TP53* or *NPM1* mutations (**Table S5, Fig. S5**).

### Number and VAF of MR-GMs do not impact outcome

Aside from *RUNX1*, the most commonly mutated MR genes in the AML-MR cohort were *ASXL1* (35%), *SRSF2* (33%), *BCOR* (20%), and *STAG2* (18%) (**Table S6**). The number of canonical MR-GM per case ranged from 1 to 5 (VAF range: 2% to 97%), and *RUNX1* mutations per case ranged from 1 to 6 (VAF range: from 2% to 95%). Median VAFs and the IQR of individual MR-GM and *RUNX1* mutations are shown in **Table S6**.

To determine whether mutation burden or clone size within AML-MR has prognostic relevance, we performed Kaplan-Meier analysis stratified by (1) total number of MR mutations (1 vs ≥2) and (2) highest VAF among all MR-GM per case. Among all 3 mutation-defined AML-MR subgroups (MR gene, RUNX1-MR, and RUNX1), the presence of two or more mutations did not confer worse OS than a single mutation (**Fig. 2A-B**). Similarly, a highest MR-GM VAF ≥10% did not portend worse survival compared to cases with smaller clone sizes (highest VAF <10%) (**Fig. 2C-D**) when classified by either WHO5th (median OS: 19 vs 36 months; *p*=0.3) or ICC (median OS: 19 vs 36 months; *p*=0.14). A VAF cut-off of 20% and 30% were also used, which did not show significant association with an inferior survival with larger clone size (data not shown). These findings were consistent across all four classification algorithms.

**Figure 2:**
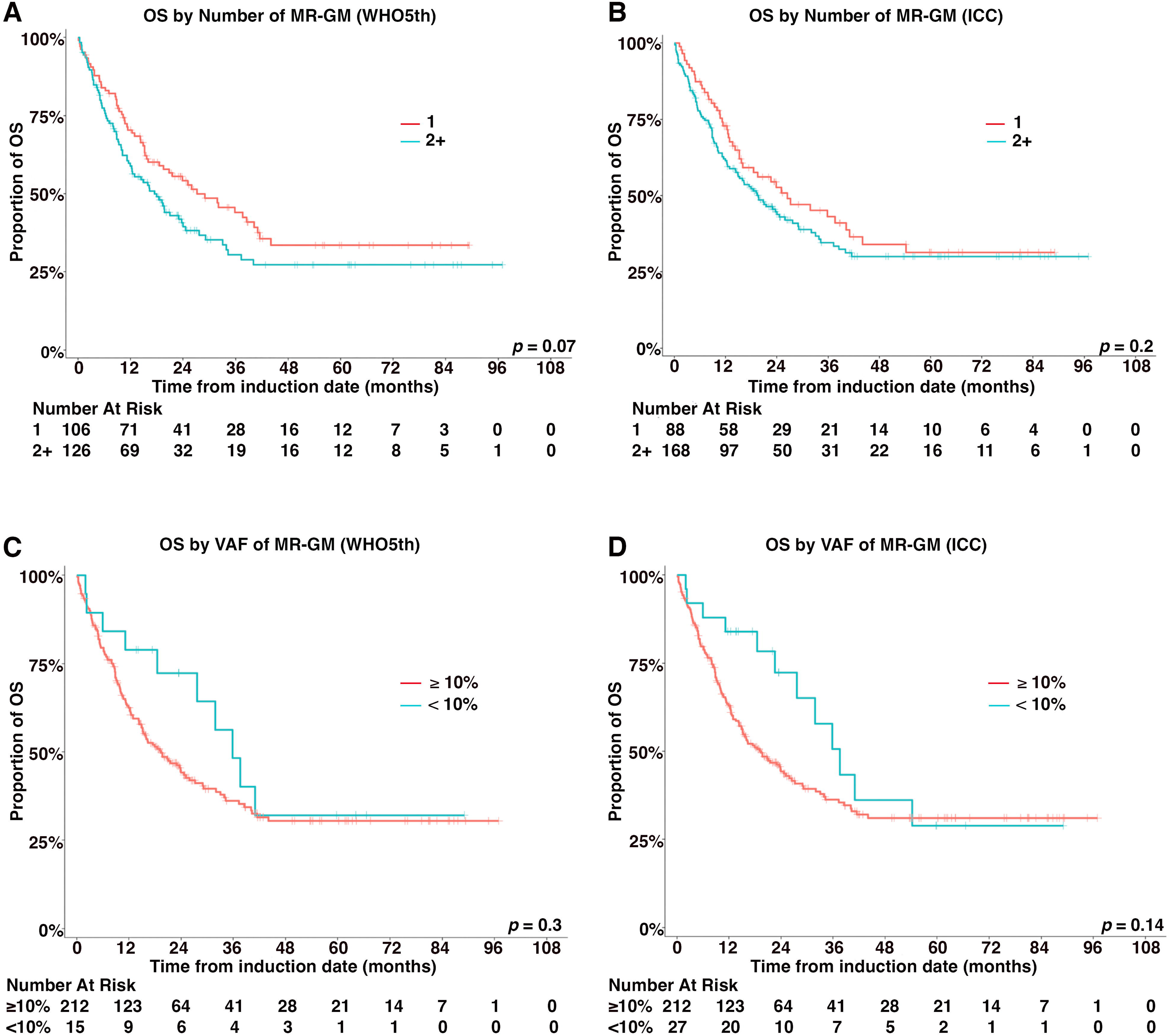
Impact of MR gene mutation number and variant allele frequency (VAF) on AML-MR outcomes. **(A–B)** Overall survival of AML-MR patients stratified by the number of MR-GM (1 vs. ≥2) as defined by WHO5th **(A)** and ICC **(B)**. **(C–D)** Overall survival of AML-MR patients stratified by the highest variant allele frequency (VAF) of any MR-GM (<10% vs. ≥10%) as defined by WHO5th **(C)** and ICC **(D)**.

### AML ontogeny does not independently impact prognosis in AML-MR

To evaluate the independent prognostic impact of clinical ontogeny in AML-MR, we compared OS among de novo, therapy-related, and secondary AML within the AML-MR group by multivariable analysis (excluding *TP53* mutated cases). After adjusting for age, sex, CG risk groups, ECOG PS, ASCT, and molecular groups, clinical ontogeny did not independently predict OS within AML-MR (*p*=0.3) (**Table 2**). In subsequent subgroup multivariable analyses stratified on treatment intensity, ASCT remained the only clinicopathologic variable with independent prognostic value among each of the 4 MR subgroups, regardless of other characteristics (**Table 2; Tables S7-8**). These data suggest that the previously reported adverse prognostic effect of secondary ontogeny is largely attributable to its association with the molecular features that define AML-MR, rather than representing an independent prognostic driver.

**Table 2:**
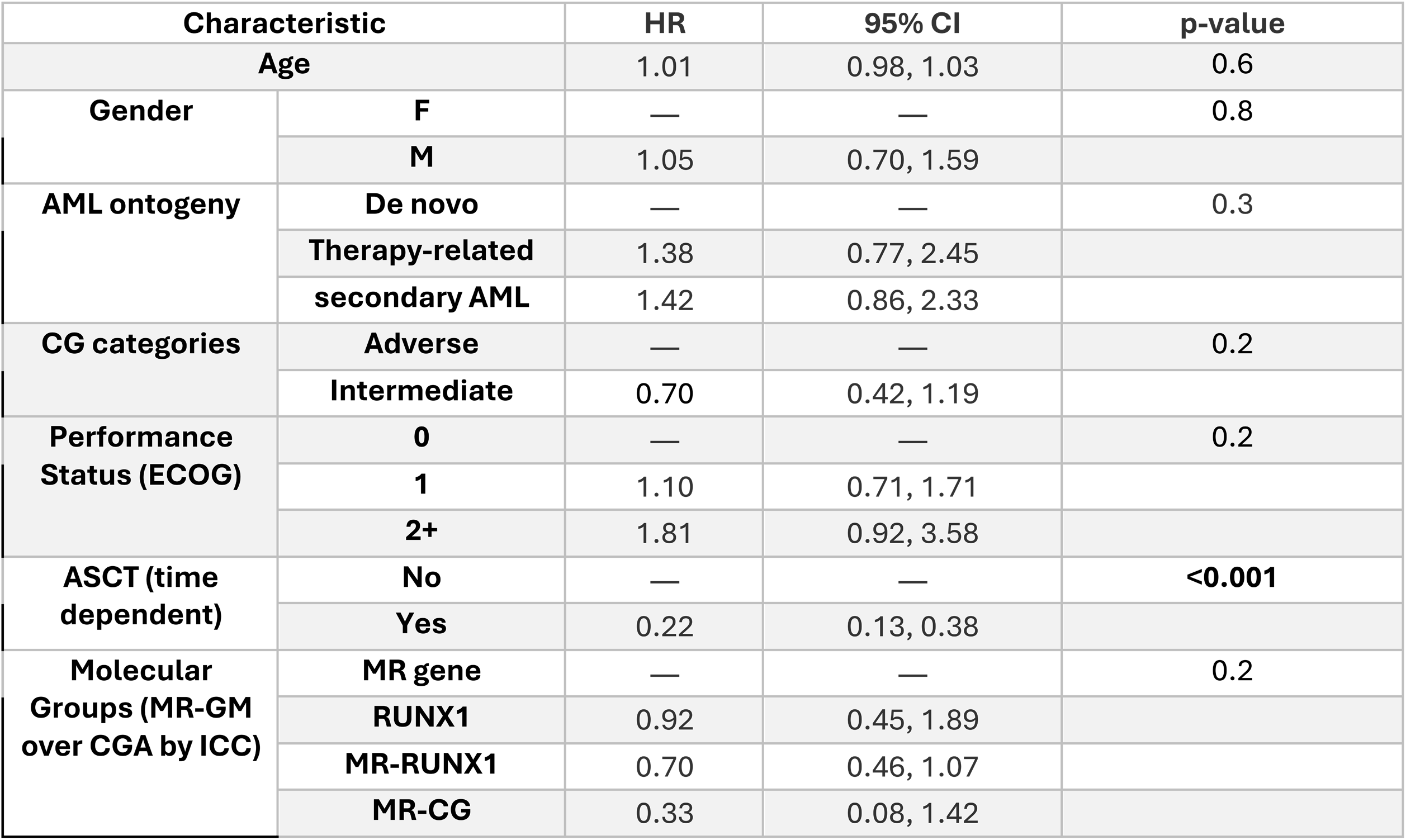
Multivariable analysis of AML-MR patients.

Since cases with a history of MDS or MDS/MPN but lacking MR-GM and/or MR-CGA were intentionally excluded from AML-MR and reclassified as AML NOS, we assessed the prognostic impact of ontogeny within this group. A prior history of MDS or MDS/MPN was not associated with inferior outcomes (**Fig. S6**). Although therapy-related AML was associated with shorter OS on univariate analysis (median OS: 27, unreached, and unreached months for therapy-related, secondary, and de novo AML, respectively), this association did not retain statistical significance on multivariate analysis (**Table S9**), suggesting that inferior outcomes in this subgroup may be driven by underlying comorbidities (as reflected by performance status) rather than therapy-related ontogeny per se.

### Trisomy 8 and del(20q) as ICC-specific MR-CGA

Trisomy 8 and del(20q) are recognized as MR-CGA by ICC but not by WHO5th. To investigate the prognostic significance of these abnormalities, we compared OS in AML-MR patients carrying only trisomy 8 and/or del(20q) (in the absence of other MR-GM or MR-CGA) to that of other AML-MR patients. Although case number was limited (n=8; 3 de novo AML, 5 secondary or therapy-related AML), patients with trisomy 8 and/or del(20q) demonstrated superior OS (median OS: unreached) in the absence of other MR-GM and/or CGA, compared to other AML-MR cases (median OS: 21 months) (*p=*0.04) (**Fig. 3A**). These findings suggest that isolated trisomy 8 and del(20q) may not confer the same adverse prognosis as other MR-CGA, and their inclusion as AML-MR defining abnormalities by ICC warrants further investigation in larger cohorts. Of note, the number of AML-MR cases harboring isolated del(11q) (n=1) or del(13)/(13q) (n=0), the two WHO5th-specific MR-CGA, was very small, precluding meaningful statistical analysis.

**Figure 3:**
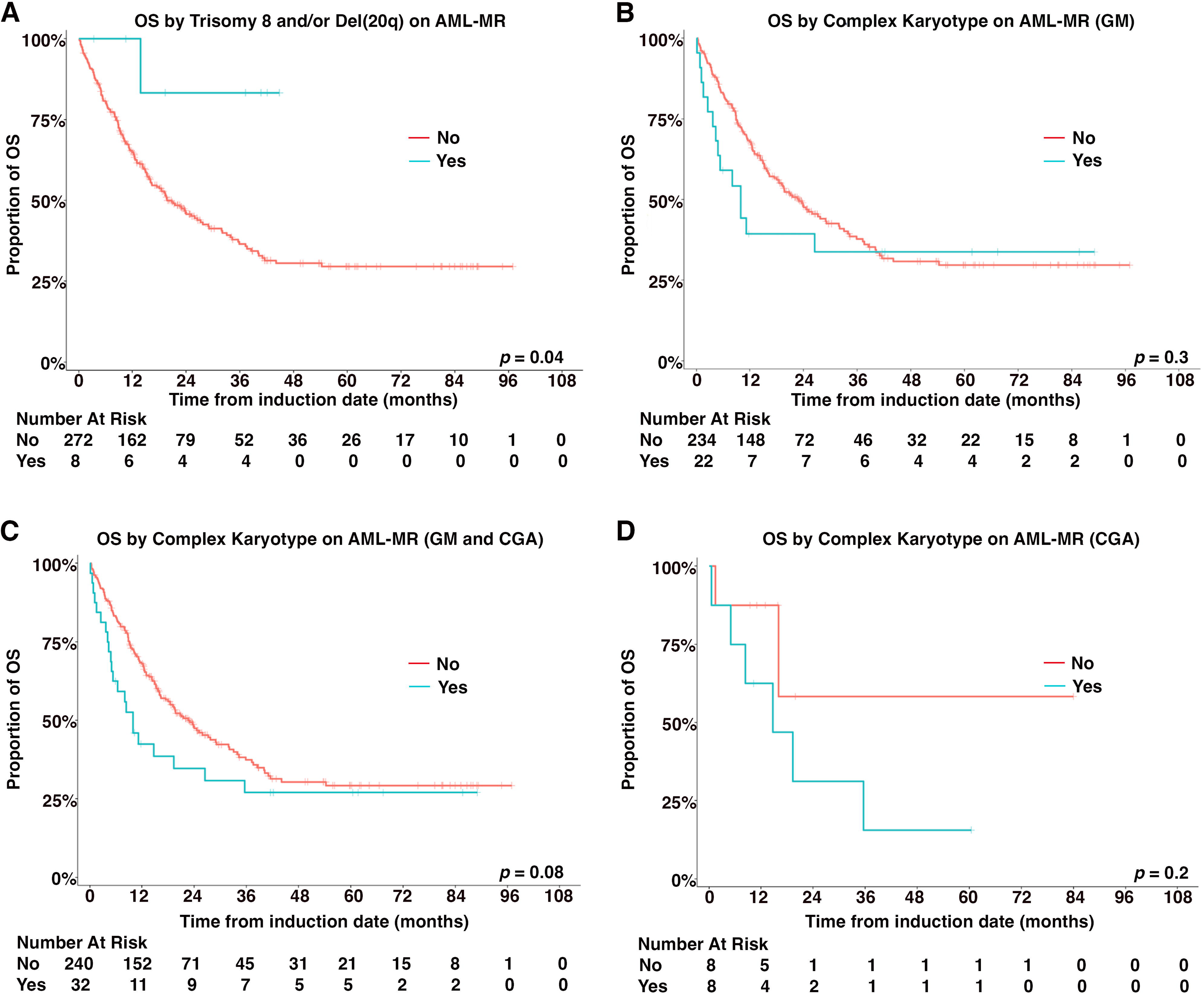
Impact of trisomy 8, del(20q), and complex karyotype (CK) on AML-MR outcomes. **(A)** Overall survival comparing AML-MR patients with isolated trisomy 8 and/or del(20q) (no concurrent MR-GM or other MR-CGA) versus all other AML-MR patients. **(B–C)** Overall survival stratified by the presence or absence of CK in AML-MR patients defined by MR-GM alone per ICC **(B)**, and by both MR-GM and MR-CGA (excluding trisomy 8 and del(20q) per ICC **(C). (D).** Overall survival of AML-MR patients defined by MR-CGA without MR-GM. Patients with isolated trisomy 8 and/or del(20q) are excluded.

### Complex karyotype without *TP53* mutations does not confer additional adverse risk in AML-MR

To determine whether CK, independent of *TP53* mutations, worsens prognosis in AML-MR, we compared OS in AML-MR patients with CK (without concurrent *TP53* mutation) to those with other MR-CGA or MR-GM without CK. No statistically significant difference in OS was observed between these two groups when AML-MR was defined by either GM (median OS: 9.9 vs 23 months; *p*=0.3) or by both GM and CGA (median OS: 9.9 vs 24 months; *p*=0.08, **Fig. 3B-C**). These findings indicate that CK, when not accompanied by *TP53* mutation, does not carry incremental adverse prognostic significance within AML-MR and should be considered equivalent to other MR-CGA for classification purposes. There were a small number of AML-MR patients carrying only MR-CGA without concurrent MR-GM mutations. The presence of CK did not confer worse outcomes compared to other MR-CGA cases (*p*=0.2, **Fig. 3D**).

### *ASXL1* and *EZH2* mutations drive adverse prognosis in AML-MR

To determine whether individual GM contributes differentially to outcomes, we performed gene-level survival analysis across the entire cohort. Significantly shorter OS was associated with mutations of *TP53*, *ASXL1*, and *EZH2*, while significantly longer survival was associated with mutations of *NPM1* and *IDH2* (**Table S10**).

To assess the contribution of individual mutations specifically within AML-MR, we performed the same gene-level survival analysis restricted to AML-MR cases (**Fig. 4A-B, S7**). Although no single GM reached statistical significance for OS within this subgroup, likely reflecting limited statistical power of smaller sample size, a consistent trend toward inferior OS was observed for *ASXL1* or *EZH2* mutated cases (**Fig. 4A-B**). To further assess the combinatorial impact of these frequently co-occurring mutations, we performed correlation analysis (**Table S11, Fig. 4C**), which revealed significant positive correlations between *ASXL1* and *EZH2*, *SRSF2*, *ZRSR2* or *STAG2*; between *SRSF2* and *STAG2*; and between *ZRSR2* and *EZH2*. Significant negative correlations were observed between *ASXL1* and *BCOR* or *SF3B1*; and between *SRSF2* and *EZH2, SF3B1* or *U2AF1*. Given the trend towards worse OS for *ASXL1* and *EZH2* mutations individually and their more frequent co-occurrence, we performed survival analysis for pooled cases with *ASXL1* and/or *EZH2* mutations compared to the remaining AML-MR cases. Inferior OS was observed for the pooled *ASXL1* and/or *EZH2* group within AML-MR (median OS: 25 vs 19 months; *p*=0.045, **Fig. 4D**).

**Figure 4:**
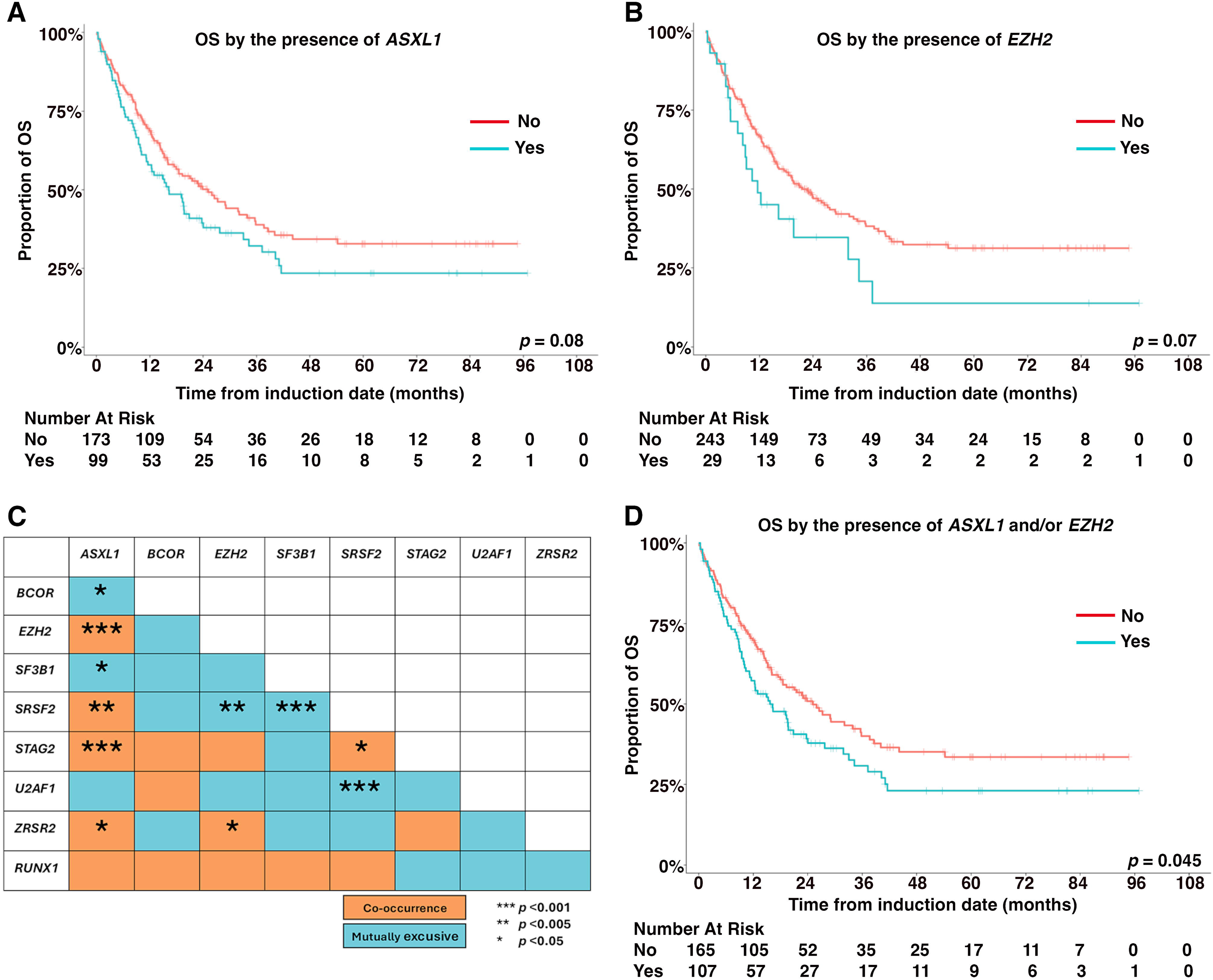
Co-mutation patterns and prognostic impact of *ASXL1* and *EZH2* mutations in AML-MR. **(A–B)** Overall survival of AML-MR patients stratified by the presence or absence of *ASXL1* **(A)** and *EZH2* **(B)** mutations. **(C)** Co-mutation correlation matrix of MR genes, including *RUNX1*, among AML-MR patients. **(D)** Overall survival of AML-MR patients stratified by the presence or absence of *ASXL1* and/or *EZH2* mutations. Patients with isolated trisomy 8 and/or del(20q) (without MR-GM or other MR-CGA) are excluded.

### *TP53*-mutated AML is prognostically distinct from AML-MR

*TP53*-mutated AML demonstrated the worst OS of all AML groups in this cohort, with a median OS of 6.9 months. Outcomes were uniformly poor regardless of VAF ≥10% or <10% (median OS: 6.9 vs 19 months; *p*=0.5), regardless of multi-hit versus single-hit allelic status (median OS: 5.8 vs 7.3 months; *p*=0.5), and regardless of the presence versus absence of complex karyotype (median OS: 6.9 vs 11 months; *p*=0.6, **Fig. 5A-C**). Within the AML-MR framework, the presence of concurrent MR-GMs in *TP53*-mutated AML did not significantly alter OS when *RUNX1* as GMs was either excluded (median OS: 6.9 vs 6.6 months, *p*=0.2) or included (median OS: 6.9 vs 7.3 months, *p*=0.8, **Fig. 5D-E**), confirming that *TP53* mutation, not MR-GM co-occurrence, is the dominant prognostic driver in these cases. These data support the ICC approach of classifying *TP53*-mutated AML (including single-hit *TP53*) as a separate entity superseding AML-MR.

**Figure 5:**
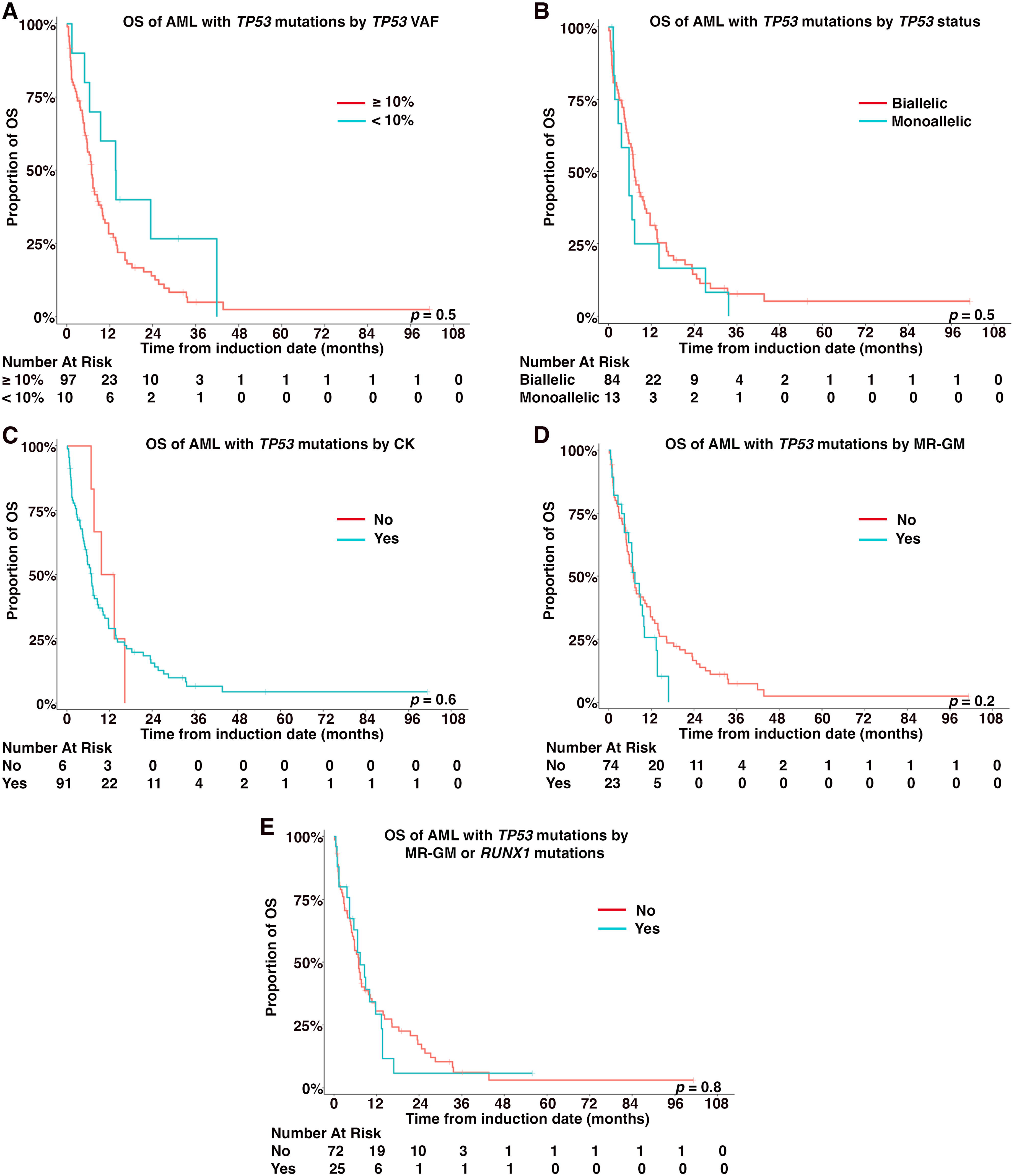
Prognostic impact of *TP53* VAF, allelic status, complex karyotype, and co-occurring MR-GM in *TP53*-mutated AML. **(A–B)** Overall survival stratified by the highest *TP53* VAF (<10% vs. ≥10%) **(A)** and allelic status (monoallelic vs. biallelic) **(B)**. **(C)** Overall survival stratified by the presence or absence of CK. (**D–E**) Overall survival stratified by the presence or absence of co-occurring MR-GM as defined by WHO5th **(D)**, and by co-occurring MR-GM including *RUNX1* mutations **(E)**.

## Discussion

Our study interrogates key areas of divergence between the WHO5th and ICC classification systems and provides data supporting a simplified, unified approach to AML-MR classification. AML-MR, whether defined by MR-GM, MR-CGA, or a combination of both, forms a discrete prognostic group with inferior outcomes compared to most other AML subtypes, with *TP53*-mutated AML and *EVI*1-rearranged AML demonstrating worse survival. This finding was robust across all 4 classification algorithms, regardless of whether WHO5th or ICC criteria were applied or whether MR-GM or MR-CGA were prioritized, underscoring the biological and clinical validity of AML-MR as a unified entity.

The inclusion of *RUNX1* mutations as MR-GMs remains one of the most consequential points of divergence between the two systems. Our data provide several independent lines of evidence against classifying isolated *RUNX1* mutations equivalently to the other 8 canonical MR mutations.

First, isolated *RUNX1* mutations were rarely associated with AMN a hallmark of AML-MR biology and were instead more frequently observed following prior cytotoxic exposure. Second, patients with isolated *RUNX1* mutations demonstrated longer OS compared to other MR subgroups, although this did not reach statistical significance, likely due to the low frequency of isolated *RUNX1* mutations in the absence of other MR-GMs or CGA. In addition, *RUNX1*-mutated AML frequently exhibits unique biological features including plasmacytoid dendritic cell differentiation and lineage infidelity, not characteristic of other canonical MR mutations.^22,23^ These findings support the WHO5th approach of excluding isolated *RUNX1* mutations from AML-MR. This exclusion carries minimal practical consequence, as approximately 90% of *RUNX1*-mutated cases co-occurred with other MR-MG or MR-CGA in our series and were appropriately captured by the WHO5th classification without requiring *RUNX1* as an independent criterion. Furthermore, our prior work demonstrated that *RUNX1* mutations in mixed phenotype acute leukemia (MPAL) do not confer inferior outcomes and therefore should not serve as grounds for reclassification as AML.^23^ Removing *RUNX1* from the MR-GM criteria would thus sharpen the diagnostic boundary between MPAL and AML, reducing potential misclassification and ensuring more biologically coherent disease categorization.

Neither WHO5th nor ICC specifies a VAF threshold for MR-GM, although ICC suggests a minimum VAF of 10%.^24^ In our cohort, OS was independent of both the highest MR-GM VAF and the total number of MR-GMs, consistent across multiple VAF cutoffs and all four classification algorithms. These findings are consistent with Esteve et al,^25^ who found no survival differences by mutation number using a similar canonical mutation framework, and contrast with studies employing expanded, non-canonical mutation definitions that reported worse outcomes with increasing mutation burden,^13^ suggesting that apparent mutation burden effects in those studies may reflect inclusion of biologically distinct non-canonical mutations. Our data support any single canonical MR-GM, regardless of VAF, as sufficient to identify adverse-risk AML-MR.

While all 8 canonical MR-GMs are currently treated as prognostically equivalent, emerging evidence suggests individual mutations may confer variable risk. Recent studies have identified *ASXL1, RUNX1, SF3B1,* and *U2AF1* as associated with relatively worse outcomes within AML-MR.^26,27^ Our data corroborate these findings, demonstrating a trend toward inferior OS in cases harboring *ASXL1* or *EZH2* mutations individually, and a statistically significant adverse impact when both mutations are considered together. This combinatorial effect is biologically plausible given the functional interconnection of *ASXL1* and *EZH2* through Polycomb Repressive Complex 2 (PRC2),^28^ where loss-of-function mutations converge on impaired H3K27 trimethylation and dysregulated epigenetic silencing. These findings suggest that specific mutations, particularly *ASXL1* and *EZH2*, may disproportionately drive the adverse prognosis of AML-MR as a group and support investigation of *ASXL1* and/or *EZH2* mutations as a basis for further molecular risk sub-stratification within AML-MR. ^29–31^

Trisomy 8 and del(20q) are recognized as MR-CGA by ICC but not WHO5th. Cases defined exclusively by these abnormalities demonstrated significantly superior OS compared to other AML-MR cases, arguing against their inclusion as AML-MR-defining features. These findings are concordant with Zhou et al, who similarly observed improved overall survival in cases defined exclusively by ICC-specific MR-CGAs, an effect likely driven by trisomy 8 and/or del(20q) ^32^ and the study of Kwon et al.^33^ Although limited by small sample sizes, these data warrant further investigation in larger cohorts before retaining these abnormalities as AML-MR-defining criteria. Conversely, the prognostic significance of CK within AML-MR, independent of *TP53* mutations, has been uncertain. Our data demonstrate that CK in AML-MR, when unaccompanied by *TP53* mutation, does not confer additional adverse prognosis beyond that of other MR-CGA, consistent with prior observations that the dismal prognosis traditionally ascribed to CK is largely driven by co-occurring *TP53* mutations. We therefore propose that CK, in the absence of *TP53* mutation, should be treated equivalently to other MR-CGA for classification and risk-stratification purposes.

AML ontogeny is treated as diagnostic qualifiers in ICC, but as a primary defining criterion for AML-MR in WHO5th. Prior studies have yielded conflicting results regarding its independent prognostic value,^15,16^ and performance status represents one potential confounder that has not been consistently accounted for. In our study, although ontogeny was a significant prognostic factor of the entire cohort, it lost statistical significance in multivariable analysis that included performance status in AML-MR patients, suggesting that its prognostic effect is largely attributable to its association with the intrinsic molecular features defining AML-MR rather than representing an independent driver. These data support defining AML-MR on the basis of GM and CGA rather than clinical history, consistent with the ICC position that ontogeny serves as a qualifier rather than a primary disease-defining criterion. Furthermore, ontogeny did not retain statistical significance in the AML NOS group on multivariate analysis, suggesting that its impact on outcome may be driven by confounding factors, such as underlying comorbidities, rather than representing an independent prognostic variable.

*TP53*-mutated AML demonstrated universally poor outcomes in our cohort regardless of allelic status, karyotype complexity, or co-occurring MR-GM, confirming its designation as a distinct entity separate from AML-MR. These findings support the ICC hierarchical classification in which *TP53* mutation (monoallelic or bi-allelic) supersedes AML-MR criteria and are consistent with the ELN 2022 classification of all *TP53*-mutated AML as adverse risk.

Key limitations include retrospective design and tertiary referral center setting, which may limit generalizability. Treatment heterogeneity was mitigated by stratifying analyses by treatment intensity. The use of different NGS platforms may introduce differences in mutation detection sensitivity. Several subgroup analyses, particularly for isolated *RUNX1* mutations, trisomy 8/del(20q), and individual MR-GM comparisons, were limited by small sample sizes and require validation in larger prospective cohorts. Despite these limitations, comprehensive cohort annotation and consistency across two independent institutions strengthen confidence in our principal conclusions.

In conclusion, our data demonstrate that AML-MR constitutes a discrete adverse-risk entity on the basis of any canonical MR-GM, independent of clinical ontogeny, VAF, or mutation number (**Fig. S8**), providing meaningful evidence toward future harmonization of the WHO5th and ICC systems.

## Author Contributions

Y.L., D.M.L., R.H., and W.X. conceived the study, collected and analyzed the data, and wrote the manuscript. Y.Z. and D.M.L. interpreted the cytogenetic data. B.B., D.N., and A.D. performed and analyzed statistical studies. Y.L., D.M.L., and A.C. annotated the sequencing data. X.W. and E.S. annotated the clinical data. All the authors approved the final version of the manuscript.

## Supporting information

Supplementary Tables 1-11 and figures 1-8 will be used for the link to the file on the preprint site.

## Data Availability

Deidentified individual data may be found in a data supplement available with the online version of this article.

