## Supplementary Tables 1-11 and figures 1-8 will be used for the link to the file on the preprint site. for "Toward a unified classification of acute myeloid leukemia, myelodysplasia-related: a multicenter retrospective study"

Short title: Unifying classification of AML MR

Authors: Ying Liu[1,2,#], Derek M. Loneman[3,#], Brenden Bready[4], David Nemirovsky[4], Alexa Cohen[1], Xin Wang[5], Eytan Stein[5], Yanming Zhang[6], Andriy Derkach[4], Robert P. Hasserjian[3,*], Wenbin Xiao[1,*]

Supplemental tables 1-11

Supplemental figures 1-8

### Supplementary Table 1: Univariable analysis of all patients

| Characteristic | | HR | 95% CI | *p*-value |
|---|---|---|---|---|
| **Age** | | 1.03 | 1.02, 1.04 | **<0.001** |
| **Gender** | **F** | — | — | 0.089 |
| | **M** | 1.21 | 0.97, 1.50 | |
| **AML ontogeny** | **De novo** | — | — | **<0.001** |
| | **Therapy-related** | 2.19 | 1.69, 2.85 | |
| | **secondary AML** | 1.98 | 1.52, 2.59 | |
| **CG Risk Group** | **Favorable** | — | — | **<0.001** |
| | **Intermediate** | 3.72 | 1.65, 8.40 | |
| | **Adverse** | 8.93 | 3.94, 20.2 | |
| **CK status** | **No** | — | — | **<0.001** |
| | **Yes** | 3.17 | 2.51, 4.00 | |
| **Performance Status (ECOG)** | **0** | — | — | **<0.001** |
| | **1** | 1.95 | 1.47, 2.58 | |
| | **2+** | 4.30 | 2.83, 6.53 | |
| **Induction regimen** | **High intensity** | — | — | **<0.001** |
| | **Low intensity** | 2.05 | 1.65, 2.56 | |
| **ASCT (time dependent)** | **No** | — | — | **<0.001** |
| | **Yes** | 0.45 | 0.35, 0.57 | |

### Supplementary Table 2: Multivariable analysis of all patients

| Characteristic | | HR | 95% CI | *p*-value |
|---|---|---|---|---|
| **Age** | | 1.01 | 1.0, 1.02 | 0.3 |
| **Gender** | **F** | — | — | |
| | **M** | 1.33 | 1.01, 1.75 | **0.042** |
| **AML ontogeny** | **De novo** | — | — | **0.014** |
| | **Therapy-related** | 1.62 | 1.17, 2.26 | |
| | **secondary AML** | 1.28 | 0.89, 1.83 | |
| **CK status** | **No** | — | — | **<0.001** |
| | **Yes** | 2.03 | 1.49, 2.76 | |
| **Performance Status (ECOG)** | **0** | — | — | **0.019** |
| | **1** | 1.44 | 1.08, 1.93 | |
| | **2+** | 1.73 | 1.07, 2.81 | |
| **ASCT (time dependent** | **No** | — | — | **<0.001** |
| | **Yes** | 0.36 | 0.26, 0.51 | |

**Supplementary Table 3: Clinical and pathologic characteristics of AML-MR patients**

| Characteristics | Total numbers (n=280) | |
|---|---|---|
| Gender (%) | Female: n = 97 (35%) | |
| | Male: n = 183 (65%) | |
| Age: median (range) | 68 (18 - 91 yo) | |
| Antecedent malignancy (%) | n = 64 (23%) | |
| HSCT (%) | n = 151 (54%) | |
| Type of AML (%) | De novo | n = 160 (57%) |
| | t-AML | n = 41 (15%) |
| | secondary AML | n = 79 (28%) |
| BM blasts%: median (range) | 43 (20-98) | |
| PB blasts%: median (range) | 7 (0-29) | |
| CBC: median (range) | ANC (K/uL) | 0.6 (0-37.4) |
| | Hgb (g/dL) | 8.5 (5.9-14.8) |
| | PLT (K/uL) | 60 (4-697) |
| Performance Status (ECOG) (%) | 0 | n = 96 (47%) |
| | 1 | n = 91 (44%) |
| | 2+ | n = 19 (9.2%) |
| | unknown | n = 74 |
| Induction regimen | High intensity | n = 140 (50%) |
| | Low intensity | n = 139 (50%) |
| | unknown | n = 1 |
| ELN2022 | Favorable | n = 0 (0%） |
| | Intermediate | n = 8 (2.9%) |
| | Adverse | n = 272 (97%) |
| ELN2024 | Favorable | n = 103 (75%) |
| | Intermediate | n = 35 (25%) |
| | Adverse | n = 0 (0%) |
| | Not applicable | n = 142 |
| CG categories | Favorable | n = 0 (0%) |
| | Intermediate | n = 216 (78%) |
| | Adverse | n = 60(22%) |
| | Unknown | n = 4 |
| Complex karyotype | n = 30 (11%) | |
| Molecular groups Molecular groups (CGA by ICC2022 over MR-GM) | MR gene | n = 82 (29%) |
| | MR-RUNX1 | n = 69 (25%) |
| | RUNX1 | n = 12 (4%) |
| | MR-CG | n = 117 (42%) |
| Molecular groups Molecular groups (MR-GM by ICC2022 over CGA) | MR gene | n = 116 (41%) |
| | MR-RUNX1 | n = 116 (41%) |
| | RUNX1 | n = 24 (9%) |
| | MR-CG | n = 24 (9%) |

**Supplementary Table 4: Clinical characteristics of patients of MR-RUNX1, MR gene, and RUNX1 subgroups**

| Characteristic | | MR-RUNX1 (n=116) | MR gene (n=116) | RUNX1 (n=24) | *p*-value |
|---|---|---|---|---|---|
| **Age: median (range)** | | 69 (64-77) | 69 (61-75) | 61 (53-64) | **<0.001** |
| **Gender: number (%)** | **F** | 30 (26%) | 36 (31%) | 15 (63%) | **0.004** |
| | **M** | 86 (74%) | 80 (69%) | 9 (37%) | |
| **AML ontogeny: number (%)** | **De novo** | 60 (52%) | 72 (62%) | 15 (63%) | **0.002** |
| | **Therapy-related** | 13 (**11%**) | 12 (**10%**) | 8 (**33%**) | |
| | **Secondary AML** | 43 (**37%**) | 32 (**28%**) | 1 (**4%**) | |
| **CG Risk Group: number (%)** | **Adverse** | 19 (17%) | 20 (17%) | 9 (37%) | 0.09 |
| | **Intermediate** | 94 (83%) | 96 (83%) | 15 (63%) | |
| **CK: number (%)** | | 7 (6%) | 10 (9%) | 5 (21%) | 0.09 |
| **Performance Status (ECOG): number (%)** | **0** | 35 (42%) | 42 (47%) | 11 (52%) | 0.6 |
| | **1** | 37 (45%) | 42 (47%) | 8 (38%) | |
| | **2+** | 11 (13%) | 6 (6.7%) | 1 (9.5%) | |
| **Treatment: number (%)** | **High intensity** | 56 (48%) | 61 (53%) | 19 (79%) | **0.02** |
| | **Low intensity** | 60 (52%) | 55 (47%) | 5 (21%) | |
| **ASCT: number (%)** | | 58 (50%) | 64 (55%) | 18 (75%) | 0.1 |

**Supplementary Table 5: Correlation analysis between *RUNX1* mutations and other gene mutations of entire cohort**

| | Absence of RUNX1 mutations (n=460) | Presence of RUNX1 mutations (n=155) | p-value |
|---|---|---|---|
| ***NPM1*** | 68 (15%) | 3 (1.9%) | **<0.001** |
| **bZIP *CEBPA*** | 28 (6.1%) | 5 (3.2%) | 0.172 |
| ***TP53*** | 108 (23%) | 7 (4.5%) | **<0.001** |
| ***ASXL1*** | 55 (12%) | 61 (39%) | **<0.001** |
| ***BCOR*** | 34 (7.4%) | 32 (21%) | **<0.001** |
| ***EZH2*** | 15 (3.3%) | 19 (12%) | **<0.001** |
| ***SF3B1*** | 25 (5.4%) | 18 (12%) | **0.009** |
| ***SRSF2*** | 54 (12%) | 56 (36%) | **<0.001** |
| ***STAG2*** | 30 (6.5%) | 25 (16%) | **<0.001** |
| ***U2AF1*** | 24 (5.2%) | 18 (12%) | **0.006** |
| ***ZRSR2*** | 6 (1.3%) | 6 (3.9%) | 0.084 |
| ***KRAS*** | 33 (7.2%) | 15 (9.7%) | 0.315 |
| ***NRAS*** | 77 (17%) | 30 (19%) | 0.458 |
| ***IDH1*** | 31 (6.7%) | 12 (7.7%) | 0.672 |
| ***IDH2*** | 58 (13%) | 25 (16%) | 0.267 |
| ***DDX41*** | 7 (1.5%) | 0 (0%) | 0.201 |

**Red**: positive correlation; **Blue**: negative correlation

**Supplementary Table 6: Frequencies and variance allelic frequencies of MR-GM**

| MR-GM | Frequencies (MR cohort) | Frequencies (Entire cohort) | VAF (range) | VAF (IQR) | VAF (Median) |
|---|---|---|---|---|---|
| ***RUNX1*** | 50% | 23% | 0.02 - 0.95 | 0.27 - 0.47 | 0.40 |
| ***ASXL1*** | 35% | 16% | 0.04 - 0.55 | 0.21 - 0.35 | 0.28 |
| ***BCOR*** | 20% | 9% | 0.04 - 0.94 | 0.36 - 0.74 | 0.44 |
| ***EZH2*** | 10% | 5% | 0.06 - 0.97 | 0.32 - 0.73 | 0.45 |
| ***SF3B1*** | 10% | 4% | 0.05 - 0.49 | 0.32 - 0.45 | 0.37 |
| ***SRSF2*** | 33% | 15% | 0.11 - 0.92 | 0.31 - 0.47 | 0.41 |
| ***STAG2*** | 18% | 8% | 0.10 - 0.88 | 0.17 - 0.72 | 0.42 |
| ***U2AF1*** | 13% | 6% | 0.18 - 0.49 | 0.39 - 0.42 | 0.41 |
| ***ZRSR2*** | 4% | 2% | 0.13 - 0.95 | 0.27 - 0.47 | 0.40 |

**Supplementary Table 7: Multivariable analysis of AML-MR patients treated by high intensity induction**

| Characteristic | | HR | 95% CI | p-value |
|---|---|---|---|---|
| Age | | 1.01 | 0.98, 1.03 | 0.5 |
| Gender | F | — | — | 0.8 |
| | M | 1.16 | 0.67, 2.00 | |
| AML ontogeny | De novo | — | — | 0.2 |
| | Therapy-related | 1.49 | 0.72, 3.09 | |
| | 2nd AML | 1.74 | 0.84, 3.64 | |
| CK status | No | — | — | 0.9 |
| | Yes | 1.07 | 0.43, 2.65 | |
| Performance Status (ECOG) | 0 | — | — | 0.077 |
| | 1 | 1.02 | 0.58, 1.79 | |
| | 2+ | 2.81 | 1.12, 7.00 | |
| ASCT (time dependent | No | — | — | **<0.001** |
| | Yes | 0.20 | 0.10, 0.41 | |
| Molecular Groups | MR gene | — | — | 0.052 |
| | RUNX1 | 0.94 | 0.42, 2.13 | |
| | MR-RUNX1 | 0.44 | 0.24, 0.80 | |
| | MR-CG | 0.35 | 0.08, 1.50 | |

**Supplementary Table 8: Multivariable analysis of AML-MR patients treated by low intensity induction**

| Characteristic | | HR | 95% CI | p-value |
|---|---|---|---|---|
| Age | | 0.99 | 0.95, 1.04 | 0.7 |
| Gender | F | — | — | 0.7 |
| | M | 0.88 | 0.43, 1.82 | |
| AML ontogeny | De novo | — | — | 0.6 |
| | Therapy-related | 1.58 | 0.61, 4.06 | |
| | 2nd AML | 1.34 | 0.67, 2.69 | |
| CK status | No | — | — | 0.9 |
| | Yes | 1.07 | 0.43, 2.65 | |
| Performance Status (ECOG) | 0 | — | — | 0.9 |
| | 1 | 0.93 | 0.44, 1.93 | |
| | 2+ | 1.18 | 0.41, 3.35 | |
| ASCT (time dependent | No | — | — | **<0.001** |
| | Yes | 0.19 | 0.07, 0.53 | |
| Molecular Groups | MR gene | — | — | 0.5 |
| | RUNX1 | 0.66 | 0.11, 4.10 | |
| | MR-RUNX1 | 1.13 | 0.60, 2.13 | |
| | MR-CG | 0.23 | 0.03, 1.99 | |

### Supplementary Table 9: Multivariable analysis of AML, NOS patients*

| Characteristic | | HR | 95% CI | p-value |
|---|---|---|---|---|
| **AML ontogeny** | **De novo** | — | — | 0.12 |
| | **Therapy-related** | 3.26 | 1.12, 9.52 | |
| | **2nd AML** | 1.23 | 0.38, 4.03 | |
| **Performance Status (ECOG)** | **0** | — | — | 0.14 |
| | **1** | 2.56 | 1.01, 6.47 | |
| | **2+** | 1.17 | 0.14, 9.91 | |

*AML with history of MDS or MDS/MPN or therapy-related in the absence of MR-GM and/or MR-CGA or *TP53* mutations was also included as AML NOS.

**Supplementary Table 10: Univariable Analysis of gene mutations of all cases**

| Characteristic | HR | 95% CI | p-value |
|---|---|---|---|
| ***NPM1*** | **0.61** | 0.42, 0.90 | **0.013** |
| *CEBPA* | 0.62 | 0.36, 1.06 | 0.080 |
| ***TP53*** | **3.78** | 2.97, 4.82 | **<0.001** |
| *RUNX1* | 1.14 | 0.89, 1.44 | 0.3 |
| ***ASXL1*** | **1.41** | 1.09, 1.82 | **0.011** |
| *BCOR* | 1.07 | 0.77, 1.50 | 0.7 |
| ***EZH2*** | **1.74** | 1.14, 2.65 | **0.011** |
| *SF3B1* | 1.29 | 0.88, 1.90 | 0.2 |
| *SRSF2* | 1.16 | 0.89, 1.53 | 0.3 |
| *STAG2* | 1.02 | 0.7, 1.49 | >0.9 |
| *U2AF1* | 1.40 | 0.95, 2.06 | 0.091 |
| *ZRSR2* | 1.30 | 0.61, 2.75 | 0.5 |
| *KRAS* | 1.15 | 0.78, 1.70 | 0.5 |
| *NRAS* | 1.01 | 0.76, 1.33 | >0.9 |
| *IDH1* | 0.75 | 0.47, 1.21 | 0.2 |
| ***IDH2*** | **0.45** | 0.30, 0.66 | **<0.001** |
| *DDX41* | 0.87 | 0.32, 2.34 | 0.6 |

### Supplementary Table 11: Correlation analysis of all MR-GM in AML-MR

| *p*-values for correlations | | | | | | | | |
|---|---|---|---|---|---|---|---|---|
| ***Variables*** | ***ASXL1*** | ***BCOR*** | ***EZH2*** | ***SF3B1*** | ***SRSF2*** | ***STAG2*** | ***U2AF1*** | ***ZRSR2*** |
| ***BCOR*** | **0.006** | | | | | | | |
| ***EZH2*** | **<0.001** | 0.171 | | | | | | |
| ***SF3B1*** | **0.008** | 0.538 | 0.254 | | | | | |
| ***SRSF2*** | **0.005** | 0.163 | **0.002** | **<0.001** | | | | |
| ***STAG2*** | **<0.001** | 0.488 | 0.168 | 0.145 | **0.040** | | | |
| ***U2AF1*** | 0.310 | 0.213 | 0.111 | 0.151 | **<0.001** | 0.797 | | |
| ***ZRSR2*** | **0.020** | 0.769 | **0.007** | 0.259 | 0.555 | 0.535 | 0.175 | |
| ***RUNX1*** | 0.149 | 0.634 | 0.093 | 0.457 | 0.107 | 0.790 | 0.655 | 0.960 |

**Red**: positive correlation; **Blue**: negative correlation

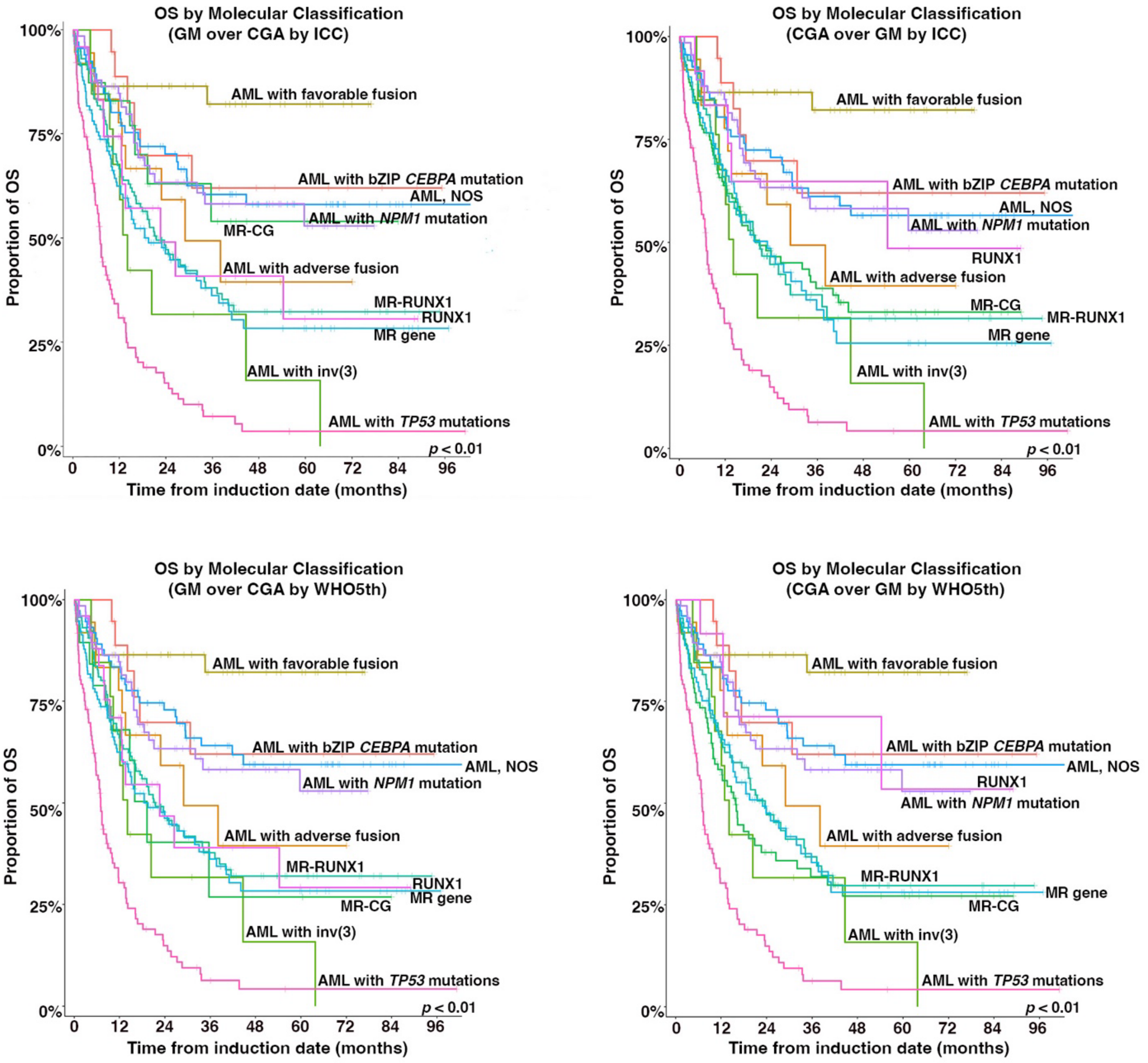


**Supplementary Figure 1**: Overall survival of all patients classified based on WHO5th and ICC2022. AML-MR is divided into 4 subgroups of MR gene, MR-RUNX1, RUNX1, MR-CG with mutation-based criteria (MR-GM) prioritized over cytogenetic criteria (MR-CGA) or verse vasa defined by either WHO5th or ICC2022.

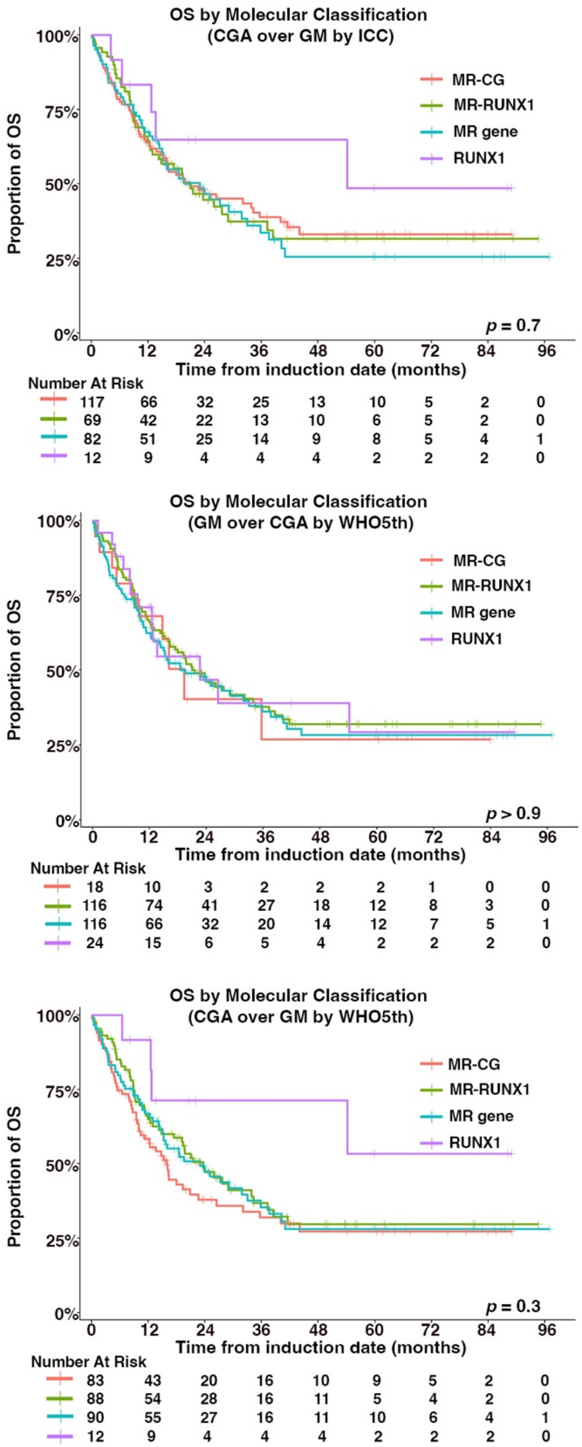


**Supplementary Figure 2**: Overall survival of AML-MR patients with mutation-based criteria (MR-GM) prioritized over cytogenetic criteria (MR-CGA) or verse vasa defined by either WHO5th or ICC2022.

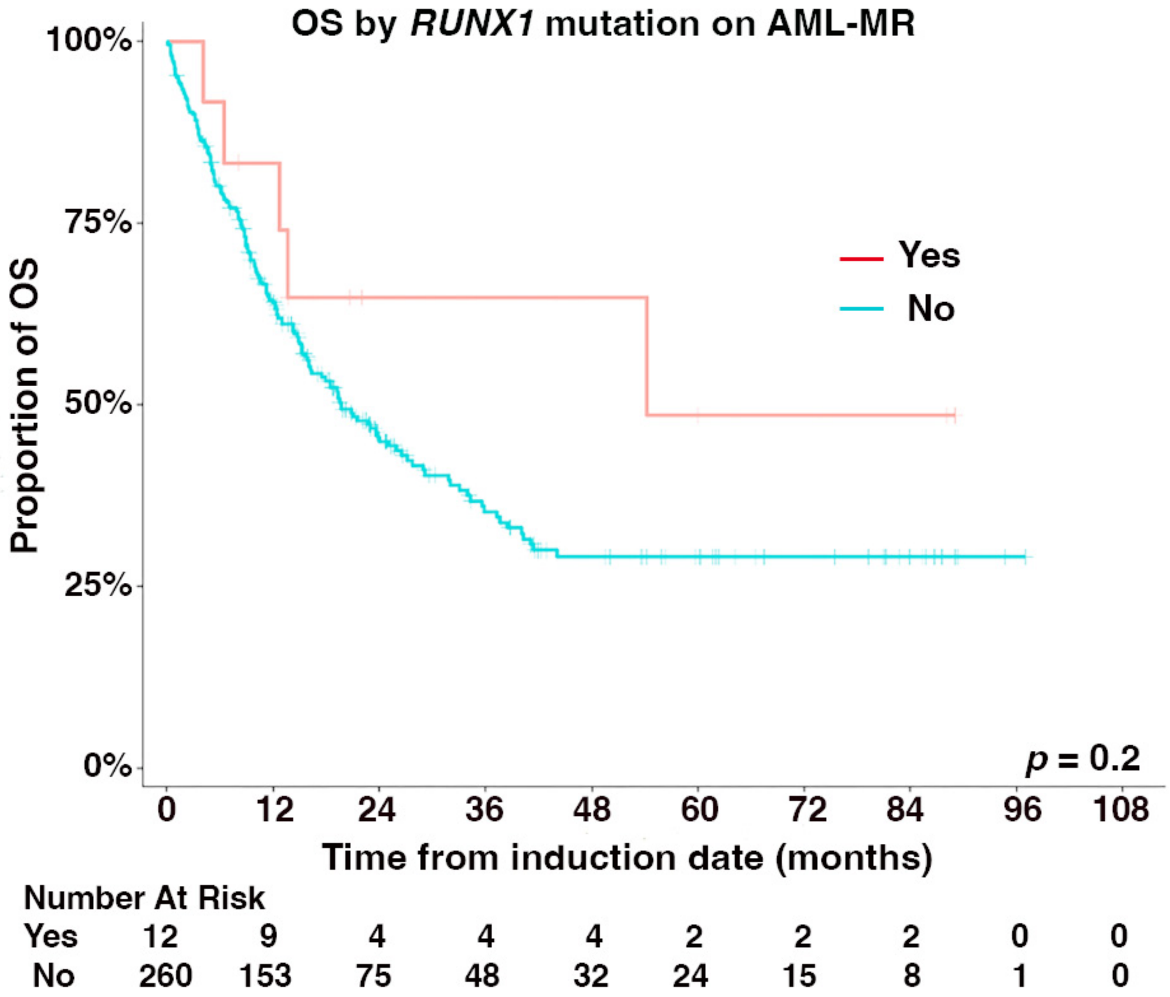


**Supplementary Figure 3:** Impacts of *RUNX1* mutations on AML-MR outcome. Overall survival of AML-MR patients with *RUNX1* mutations and no concurrent other MR-GM and CGA compared to other AML-MR patients. Patients with isolated trisomy 8 and/or del(20q) are excluded.

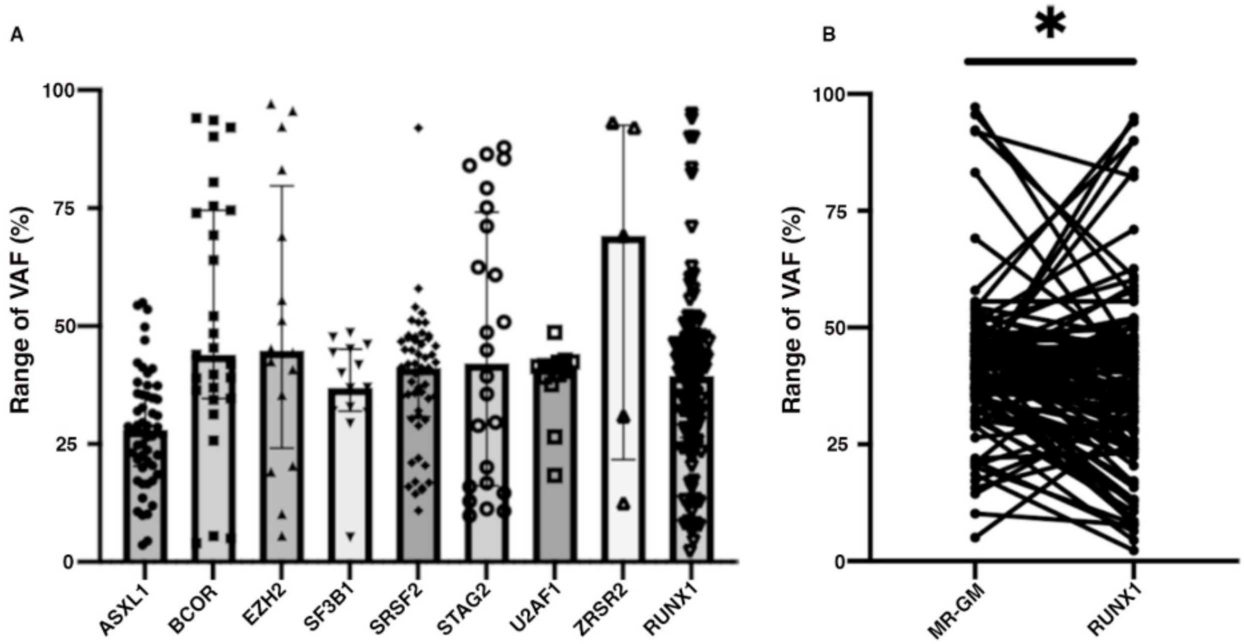


**Supplementary Figure 4: A.** Ranges of VAF of MR-GM including *RUNX1* mutations in AML-MR patients. **B.** Median VAF comparison between 8 canonical MR-GM (41.4%; IQR 35-47%) and *RUNX1* mutations (39.5%; IQR 26-47%) in subgroup of RUNX1-MR patients ($p$=0.03).

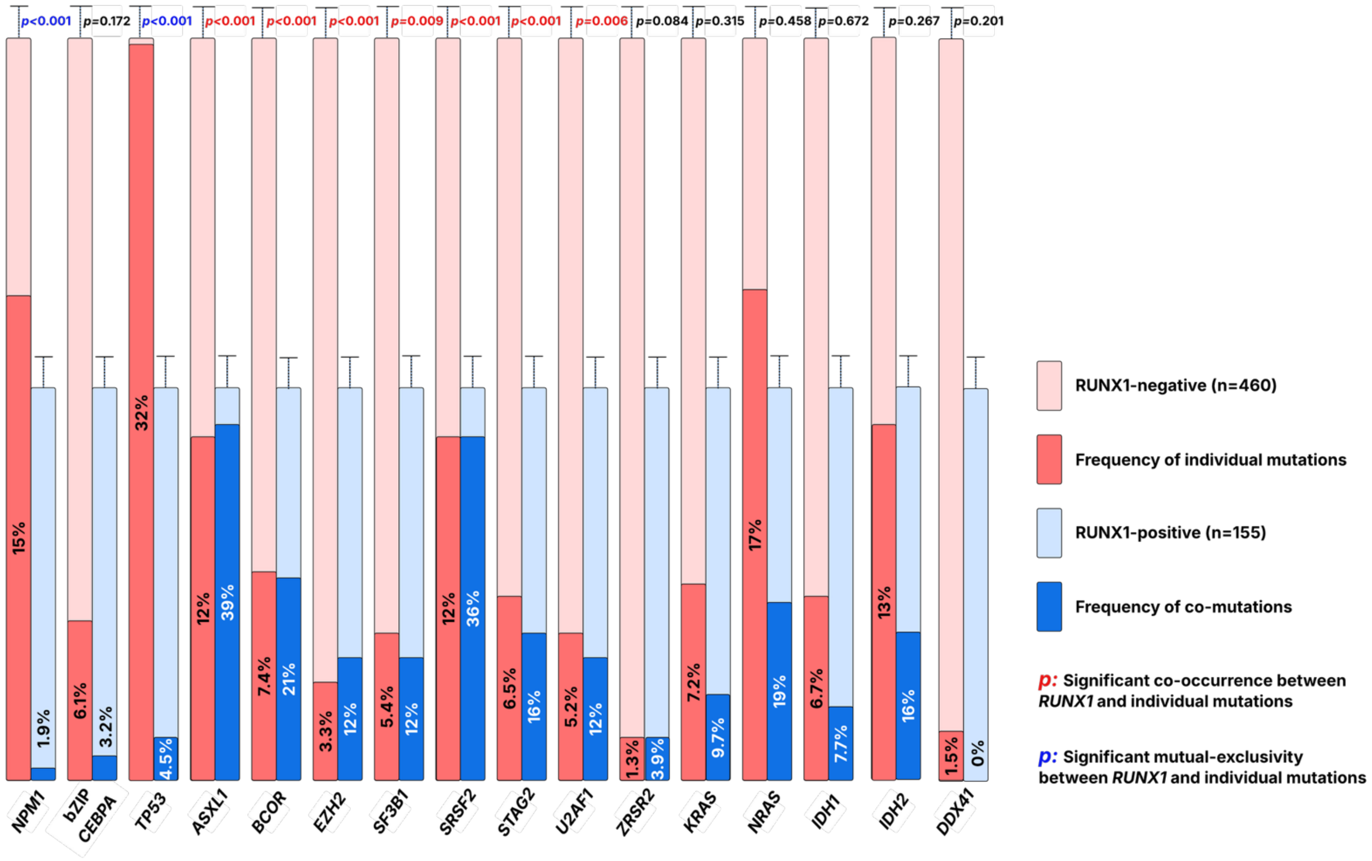


**Supplementary Figure 5:** Correlation of *RUNX1* mutations with 8 canonical MR-GM and mutations of *NPM1*, in-frame bZIP *CEBPA*, *TP53*, *KRAS*, *NRAS*, *IDH1*, *IDH2,* and *DDX41* in the entire cohort.

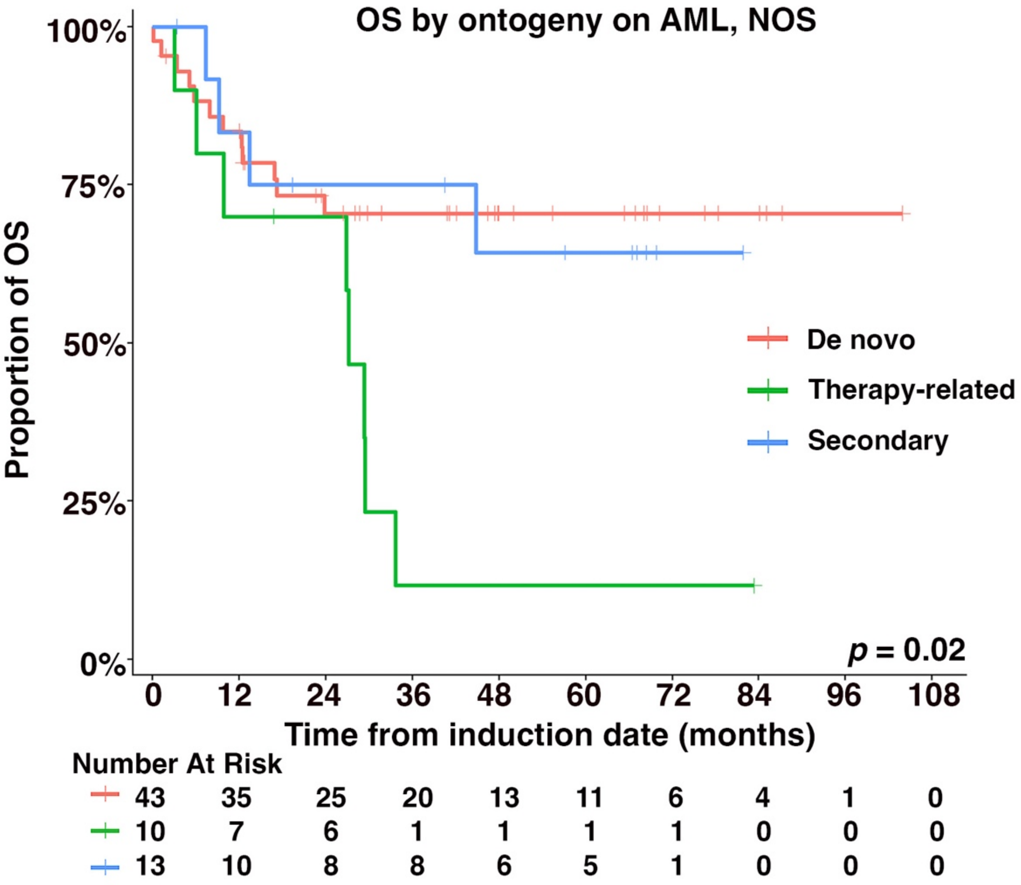


**Supplementary Figure 6:** Impacts of AML ontogeny (de novo, therapy-related, or secondary AML) on AML, NOS patients.

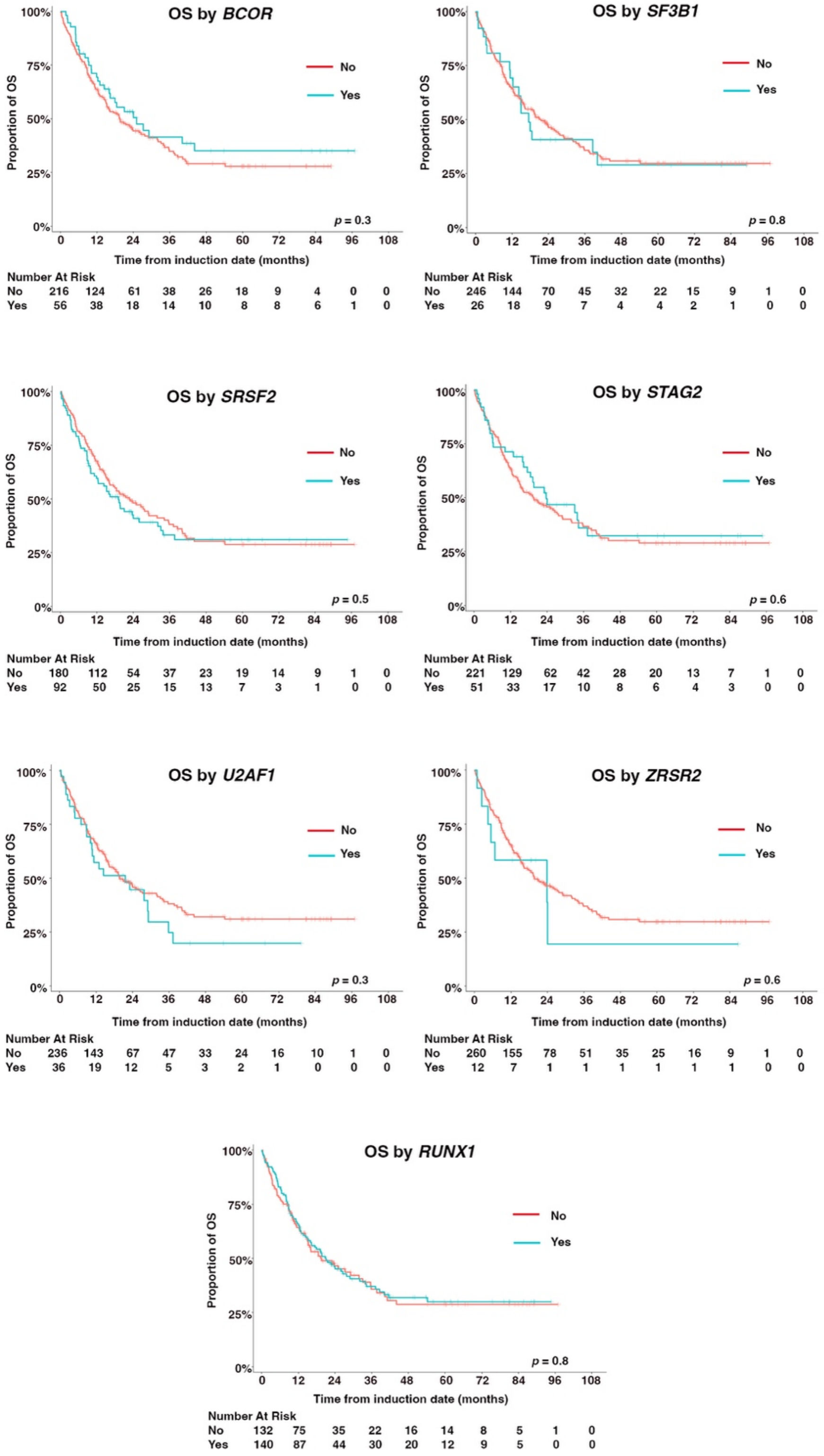


**Supplementary Figure 7:** Impacts of individual MR-GM on AML-MR outcome stratified by the presence of individual mutations. Patients with isolated trisomy 8 and/or del(20q) are excluded.

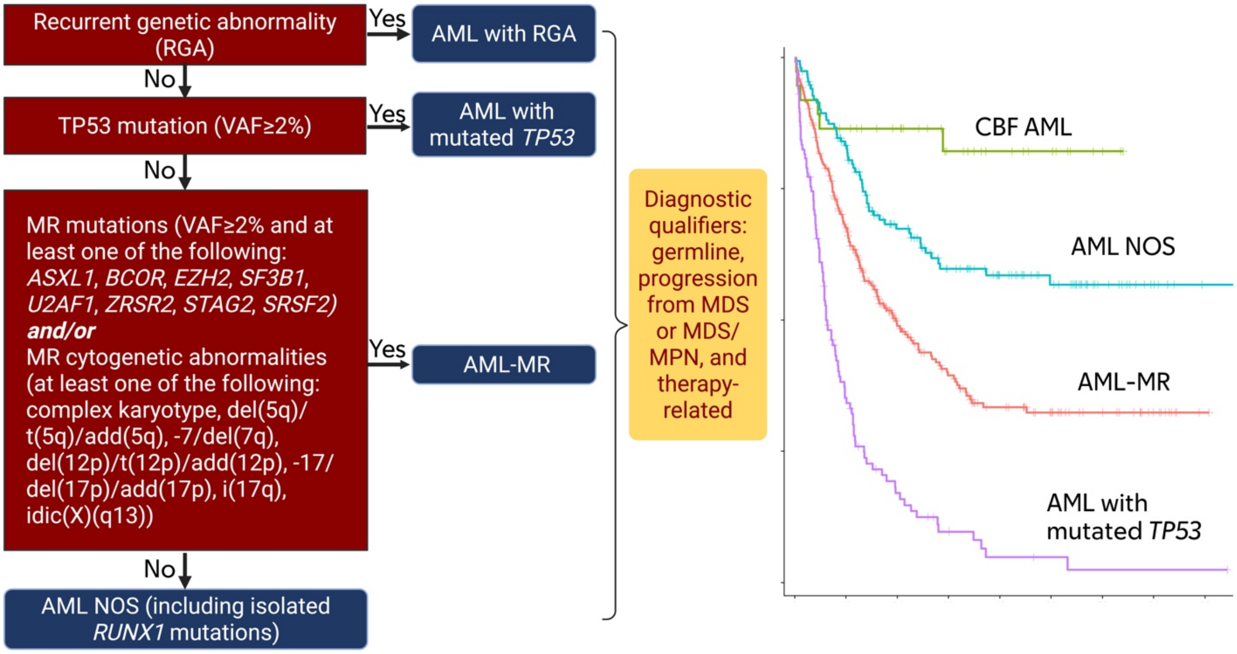


**Supplementary Figure 8:** Proposed diagnostic algorithm for AML classification. The hierarchy proceeds as follows: (1) AML with recurrent genetic abnormalities (RGA); (2) AML with mutated *TP53* (VAF ≥2%), irrespective of allelic or CK status; (3) AML-MR, defined by ≥1 of 8 MR-GM (VAF ≥2%) and/or MR-CGA; and (4) AML NOS, including cases with isolated *RUNX1* mutation without MR-GM or MR-CGA. Diagnostic qualifiers - germline predisposition, progression from MDS or MDS/MPN, and therapy-related disease - apply across all categories.
